# Muscle MRI quantifies disease progression in Amyotrophic Lateral Sclerosis

**DOI:** 10.1101/2024.04.10.24305608

**Authors:** Uros Klickovic, Luca Zampedri, Nick Zafeiropoulos, Oliver J Ziff, Christopher DJ Sinclair, Stephen J Wastling, Magda Dudziec, Jodie Allen, Karin Trimmel, Robin S Howard, Andrea Malaspina, Nikhil Sharma, Katie CL Sidle, Sachit Shah, Christian Nasel, Tarek A Yousry, Linda Greensmith, Jasper M Morrow, John S Thornton, Pietro Fratta

## Abstract

**Importance:** Quantitative, sensitive and operator-independent biomarkers of disease progression are needed to minimize the size, duration, and cost of clinical trials in amyotrophic lateral sclerosis (ALS) to allow a more effective investigation of promising therapeutic agents.

**Objective:** We assess the potential of skeletal muscle magnetic resonance imaging (MRI) as a sensitive and reliable outcome measure for future ALS clinical trials.

**Design:** In this longitudinal cohort study, muscle MRI of head-neck, upper, and lower limb regions, along with clinical and functional assessments, were acquired at three timepoints over the individual maximum observation period (iMOP) of 1 year.

**Participants:** Twenty consecutive ALS patients were recruited from a motor neuron disease clinic between 2015 and 2017, along with 16 healthy controls.

**Main outcomes and measures:** Quantitative MRI parameters CSA (cross-sectional area), VOL (muscle volume), FF (fat fraction), functional rest muscle area (fRMA) and water T2 (T_2m_) were used to assess progressive muscle degeneration and were correlated with changes in clinical parameters of disease severity (functional rating scales and myometry). Standardized response mean (SRM) was calculated for MRI outcome measures.

**Results:** Out of 20 ALS patients, 17 were available for follow-up. A significant decline in VOL of the dominant hand (rs=0.66, p<0.001) and head and neck muscles (partial η^²^=0.47, p=0.002) as well as CSA of the lower limbs (thighs: partial η^²^=0.56, p<0.001, calves: partial η^²^=0.54, p<0.001) was observed over iMOP, with highest responsiveness for progressive atrophy in hand (SRM -1.17) and leg muscles (thigh: SRM -1.09; calf: SRM -1.08). Furthermore, lower limb FF and T_2m_ increased over time, with distal leg muscles being most affected (FF: partial η^²^=0.54, p=0.002, SRM 0.81; T_2m:_ partial η^²^=0.37, p=0.01, SRM 0.74). Finally, longitudinal MRI changes showed correlation with strength in leg muscles (knee extension: r=0.77, p=0.001, 95% CI 0.46–0.91; plantar flexion: r=0.78, p<0.001, 95% CI 0.47 – 0.92), and fRMA decrease at thigh and calf level correlated with global clinical disease progression measured with ALSFRS (r=0.52, p=0.03, and r=0.68, p=0.004 respectively).

**Conclusions and Relevance:** Our findings demonstrate the effectiveness of muscle MRI as a sensitive neuroimaging biomarker of disease progression in ALS, highlighting its potential application in clinical trials.

**Key Points:** **Question:** Can quantitative muscle MRI track disease progression over time in patients with amyotrophic lateral sclerosis (ALS)?

**Findings:** Longitudinal quantitative MRI reliably detects progressive intramuscular fat infiltration, oedema and atrophy, which correlate with progressive loss of muscle strength.

**Meaning:** Muscle MRI has the potential to be utilised as a biomarker for ALS disease progression and could contribute to reduce the size and duration of clinical trials facilitating future drug developments.

## INTRODUCTION

Amyotrophic lateral sclerosis (ALS) is a rapidly progressive and fatal disorder, with a lifetime risk of approximately 1 in 350 people^1^. Upper and lower motor neurons degenerate in ALS^2^, leading to progressive bulbar and limb weakness with variable patterns of anatomical onset and spread^3^. Although promising therapeutic agents have been identified^4,5^, to date there is still no effective disease-modifying therapy, and only few agents have demonstrated effects on disease progression^4–7^ or patient survival^8^ in controlled clinical trials. Sensitive and reliable measures of disease progression would enable effective clinical trials with a shorter duration and reduced participant numbers, thereby increasing the number of therapeutic agents that could be tested.

There has been an extensive search for biomarkers to assess disease progression in motor neuron diseases (MND), which may enable new potential treatments^9–11^. One promising candidate, neurofilament levels in biofluids, has generated significant interest, although reduction in levels has not consistently been associated with clinical benefit^12^. As muscle atrophy is a direct consequence of motor neuron loss in MND, muscle MRI has been suggested as a promising imaging biomarker to sensitively quantify disease severity and progression^13^.

Quantitative muscle MRI has previously been validated to detect muscle abnormalities in various neuromuscular disorders including Duchenne muscular dystrophy^14^, inclusion body myositis (IBM)^15^ as well as in MND like spinal and bulbar muscular atrophy (SBMA)^16^. These slowly progressing neuromuscular diseases are mainly associated with increased muscle fat infiltration, which can readily be assessed by fat fraction (FF), a well-established and validated MRI parameter^17^. However, in rapidly progressing neuromuscular diseases such as ALS, FF increases are less striking, whilst loss of muscle mass is evident from clinical observation suggesting that quantification of muscle atrophy^13^ may be of greater clinical relevance.

In the present study, we applied quantitative muscle MRI of the upper and lower limbs as well as the head-neck region in ALS patients at 3 time points over one year. We used quantitative MRI methods to assess intramuscular fat infiltration, oedema and atrophy in slow and fast progressing ALS patients. MRI outcome measures were correlated with clinical disease severity over time, to determine the full potential of muscle MRI as a neuroimaging biomarker for future clinical trials in ALS.

## METHODS

Methods are provided in the Supplementary Material.

## RESULTS

### Participant demographics and clinical findings

The study included 20 ALS patients of which 17 were available for follow-up, along with 16 healthy controls^13^. Median disease duration at recruitment in ALS patients was 2 years (**Supplementary** Fehler! Verweisquelle konnte nicht gefunden werden.). Age (U=148.5, p=0.72), handedness (p=0.72), and sex (p=0.61) did not significantly differ between patients and controls. Body mass index (BMI) was significantly lower in ALS patients compared to controls (T=-2.9, p=0.01; **Supplementary** Fehler! Verweisquelle konnte nicht gefunden werden.). In ALS patients, there was a significant decline in mean ranks of ALSFRS-R and its subscales; ALSFRS-R total score (test statistic=-153, p<0.001), ALSFRS-R lower limb subscale (test statistic=-71, p=0.01), ALSFRS-R hand subscale (test statistic=-78, p<0.001) and ALSFRS-R bulbar subscale (test statistic=-45, p=0.004) over the individual maximum observation period (iMOP). Moreover, muscle strength of assessed muscle movements deteriorated significantly during follow-up; knee extension (T(1,16)=4.88, p<0.001), knee flexion (T(1,16)=5.38, p<0.001), dorsal extension (T(1,16)=4.97, p<0.001), plantar flexion strength (T(1,16)=3.94, p=0.001), jaw opening strength (T=4.07, p=0.001), hand grip strength (T(1,15)=4.70, p<0.001), and hand pinch strength (T(1,15)=4.78, p<0.001; **Supplementary Table 2**). Iowa-Oral-Pressure-Instrument (IOPI) measurements^18^ did not significantly change over time (T(1,12)=1.62, p=0.13; **Supplementary Table 2**).

### Muscle MRI detects widespread progressive atrophy in ALS

To investigate whether MRI could detect progressive muscle atrophy, we evaluated changes in the cross-sectional area (CSA) over time. At thigh level, CSA*_THIGH_* significantly decreased over the iMOP in ALS patients (partial η^²^=0.56; T(1,16)=4.51, p<0.001), while controls showed no change over 12 months (**Table 1**, **Figure 1A-B**). Similar effects were observed at calf level, with a significant decrease of CSA*_CALF_* in patients (partial η^²^=0.54; T(1,16)=4.30, p<0.001), with no changes in controls over time (**Table 1**, **Figure 1C-D**).

**Figure 1.**
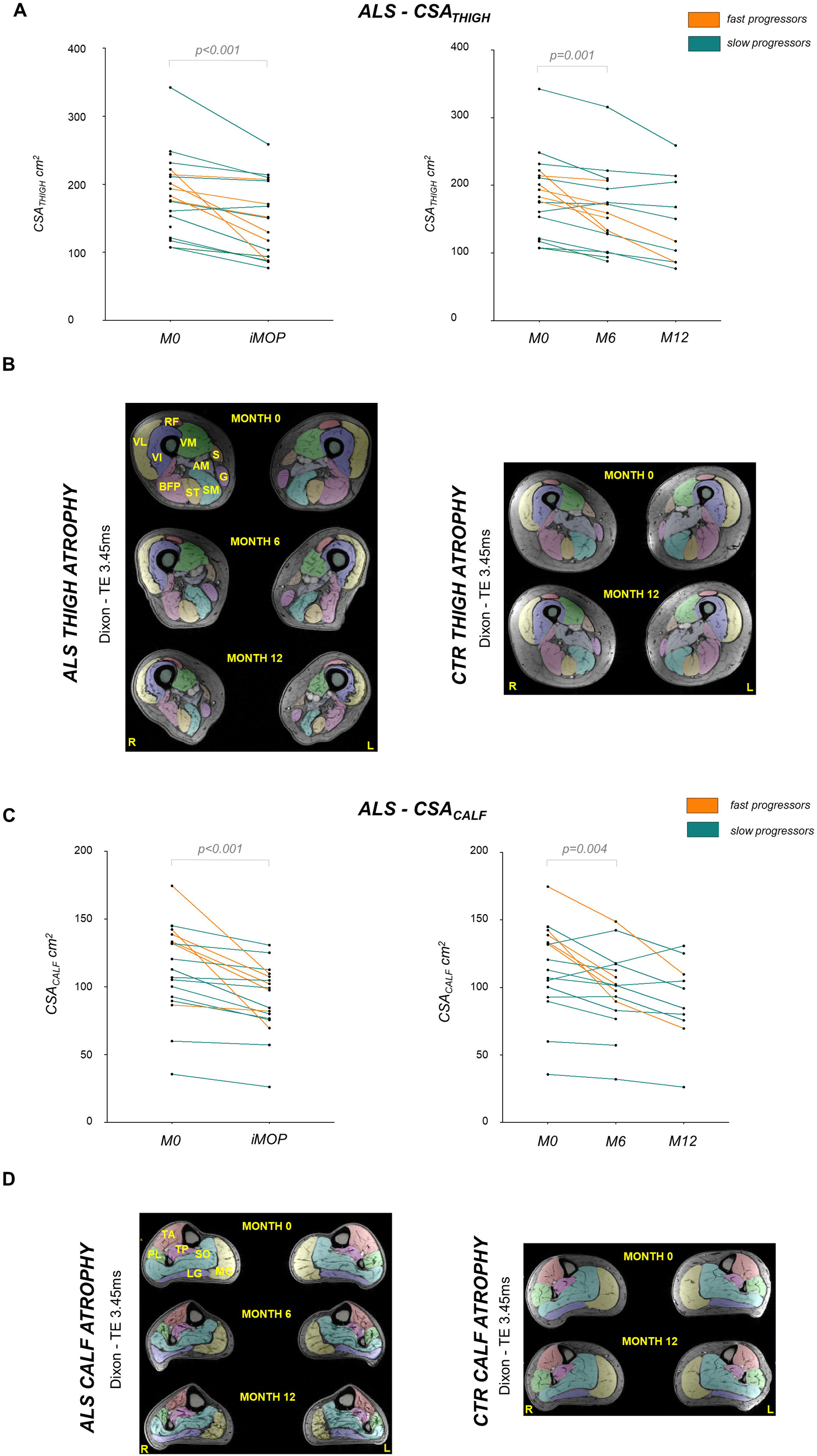
Cross-sectional area (CSA) shows progressive reduction at thigh and calf level in ALS. (**A**) Overall cross-sectional area at thigh level (CSA*_THIGH_*) in ALS patients (fast progressors: orange; slow progressors: green). Left panel: mean values at baseline (M0) and individual maximum observation period (iMOP). Right panel: individual values at baseline (M0), 6 months (M6) and 12 months (M12). CSA*_THIGH_* significantly decreased over the iMOP (T(1,16)=4.51, p<0.001) in ALS patients. Significant effects were already observed after 6 months (T(1,16)=4,11, p<0.001). **(B)** Sample axial Dixon imaging (TE=3.45ms) of thighs. Left panel: ALS patients at baseline (upper row), 6 months (middle row) and 12 months (lower row). Right panel: Controls at baseline (upper row) and 12 months (lower row). **(C)** Overall CSA at calf level (CSA*_CALF_*) in ALS patients. Left panel: mean values at baseline (M0) and iMOP. Right panel: individual values at baseline (M0), 6 months (M6) and 12 months (M12). CSA*_CALF_* significantly decreased over iMOP (T(1,16)=4.30, p<0.001). Significant effects were already observed after 6 months (T(1,15)=3.42, p=0.004). **(D)** Sample axial Dixon imaging (TE=3.45ms) of calves. Left panel: ALS patients at baseline (upper row), 6 months (middle row) and 12 months (lower row). Right panel: Controls at baseline (upper row) and 12 months (lower row). Note: ALS: amyotrophic lateral sclerosis; CSA: cross-sectional area; CSA*_THIGH_*: overall cross-sectional area of thigh muscles; CSA*_CALF_*: overall cross-sectional area of calf muscles; iMOP: individual maximum observation period; M0: baseline; M6: 6-month follow up; M12: 12-month follow-up. Muscle abbreviations: AM: adductor magus; BFP: biceps femoris posterior; G: gracilis; LG: lateral gastrocnemius; MG: medial gastrocnemius; PL: peroneus longus; RF: rectus femoris; S: sartorius; SM: semimembranosus; SO: soleus; ST: semitendinosus; TA: tibialis anterior; TP: tibialis posterior: VL: vastus lateralis; VI: vastus intermedius; VM: vastus medialis

**Table 1.** Quantitative MRI parameters in ALS patients and controls at baseline and follow-up (maximum observation period). Data are presented as mean±SD or median (IQR) according to data distribution.

| Atrophy | ALS |  |  |  | Controls |  |  |  |
| --- | --- | --- | --- | --- | --- | --- | --- | --- |
|  | Baseline | follow-up | p | 95% CI | Baseline | follow-up | p | 95% CI |
| $CSA_{CALF}$ (cm<sup>2</sup>) | 112.81±<br>33.17 | 91.24±<br>26.66 | **<0.001** | -32.22–<br>-0.93 | 146.64±<br>36.36 | 143.40±<br>32.61 | 0.13 | -2.63–<br>9.11 |
| $CSA_{THIGH}$ (cm<sup>2</sup>) | 183.33±<br>59.24 | 148.08±<br>56.23 | **<0.001** | -56.02–<br>-20.21 | 235.98±<br>59.12 | 238.59±<br>67.19 | 0.48 | -10.9–<br>21.87 |
| $VOL_{HAND}$ (cm<sup>3</sup>) | 57.04<br>(16.39) | 41.69<br>(13.98) | **<0.001** | -16.13–<br>-1.64 | 73.28±16.<br>43 | 70.56±<br>17.88 | 0.37 | -3.30–<br>1.30 |
| $VOL_T$ (cm<sup>3</sup>) | 18.33<br>(6.59) | 13.57<br>(8.17) | **<0.001** | -6.73–<br>-0.37 | 26.77±<br>6.60 | 26.86±<br>6.43 | 0.85 | -0.84–<br>1.01 |
| $VOL_{HT}$ (cm<sup>3</sup>) | 8.30<br>(2.65) | 6.45<br>(2.66) | **0.002** | -2.6–<br>-0.62 | 10.26±<br>2.91 | 10.07±<br>2.81 | 0.86 | -0.53–<br>0.63 |
| $VOL_{IO}$ (cm<sup>3</sup>) | 25.22<br>(8.67) | 18.74<br>(8.28) | **<0.001** | -6.18–<br>-0.97 | 29.60±<br>6.57 | 29.01±<br>6.36 | 0.83 | -1.35–<br>1.09 |
| $VOL_{PL}$ (cm<sup>3</sup>) | 14.24±<br>3.92 | 13.28±<br>3.70 | **0.002** | -1.51–<br>-0.40 | 15.62±<br>4.84 | 15.43±<br>4.91 | 0.38 | -0.26–<br>0.63 |
| Fat infiltration | Baseline | follow-up | p | 95% CI | Baseline | follow-up | p | 95% CI |
| $FF_{CALF}$ (%) | 3.34<br>(2.07) | 4.89<br>(7.77) | **0.002** | 0.08–<br>4.43 | 1.92<br>(1.08) | 1.92<br>(0.84) | 0.39 | -0.37–<br>0.16 |
| $FF_{THIGH}$ (%) | 2.79<br>(1.13) | 2.95<br>2.24 | **0.04** | -0.08–<br>1.2 | 1.79<br>(0.98) | 1.70<br>(1.78) | 0.49 | -0.14–<br>0.33 |
| $FF_{HAND}$ (%) | 2.22<br>(3.47) | 5.83<br>(4.81) | **<0.001** | 1.01–<br>6.08 | 2.13<br>(1.93) | 1.75<br>(1.38) | 0.45 | -1.78–<br>1.52 |
| $FF_{TONGUE}$ (%) | 8.50±<br>3.42 | 11.04±<br>6.89 | 0.25 | -1.93–<br>6.82 | 8.51±<br>5.98 | 9.44±<br>5.08 | 0.59 | -4.57–<br>2.70 |
| $fRMA_{CALF}$ (cm<sup>2</sup>) | 108.48±<br>32.67 | 85.40±<br>27.31 | **<0.001** | -35.69–<br>-12.46 | 138.82±<br>32.73 | 136.56±<br>29.40 | 0.45 | -4.04–<br>8.57 |
| $fRMA_{THIGH}$ (cm<sup>2</sup>) | 178.12±<br>57.59 | 143.02±<br>55.82 | **<0.001** | -55.51–<br>-20.32 | 231.01±<br>58.18 | 233.26±<br>65.71 | 0.51 | -10.91–<br>20.94 |
| $fRMA_{TONGUE}$ (cm<sup>2</sup>) | 25.94±<br>4.97 | 24.29±<br>5.54 | 0.52 | -4.49–<br>2.38 | 27.11±<br>4.52 | 25.48±<br>3.88 | 0.21 | -1.05–<br>4.30 |
| $T_{2m}$ | Baseline | follow-up | p | | Baseline | follow-up | p | |
| $T_{2m_{CALF}}$ (ms) | 34.61±<br>3.85 | 36.87±<br>5.48 | **0.01** | -3.90 –<br>-0.63 | 29.67±<br>0.81 | 29.50±<br>0.94 | 0.28 | -0.16 –<br>0.50 |
| $T_{2m_{THIGH}}$ (ms) | 31.43±<br>1.34 | 32.30±<br>2.01 | **0.04** | -1.67 –<br>-0.07 | 29.63±<br>1.00 | 29.54±<br>0.83 | 0.60 | -0.27 –<br>0.45 |

At hand level, atrophy was investigated by measuring muscle volume on Dixon imaging. No significant change was detected in controls, whilst overall hand muscle volume VOL*_HAND_* significantly declined over the iMOP in ALS patients (rs=0.66; test statistic=-122, p<0.001); **Table 1**, **Figure 2A**). Subanalyses of the hand included single muscle volumes of the thenar (VOL*_T_*), hypothenar (VOL*_HT_*) and interossei muscles (VOL*_IO_*); with a significant decrease of muscle volume in each subgroup (VOL*_T_*: rs=0.59, test statistic=-120, p<0.001; VOL*_HT_*: rs=0.59, test statistic=-114, p=0.002; VOL*_IO_*: rs=0.59, test statistic=-124, p<0.001, 95%CI= -6180 - -970; **Figure 2B-C**).

**Figure 2.**
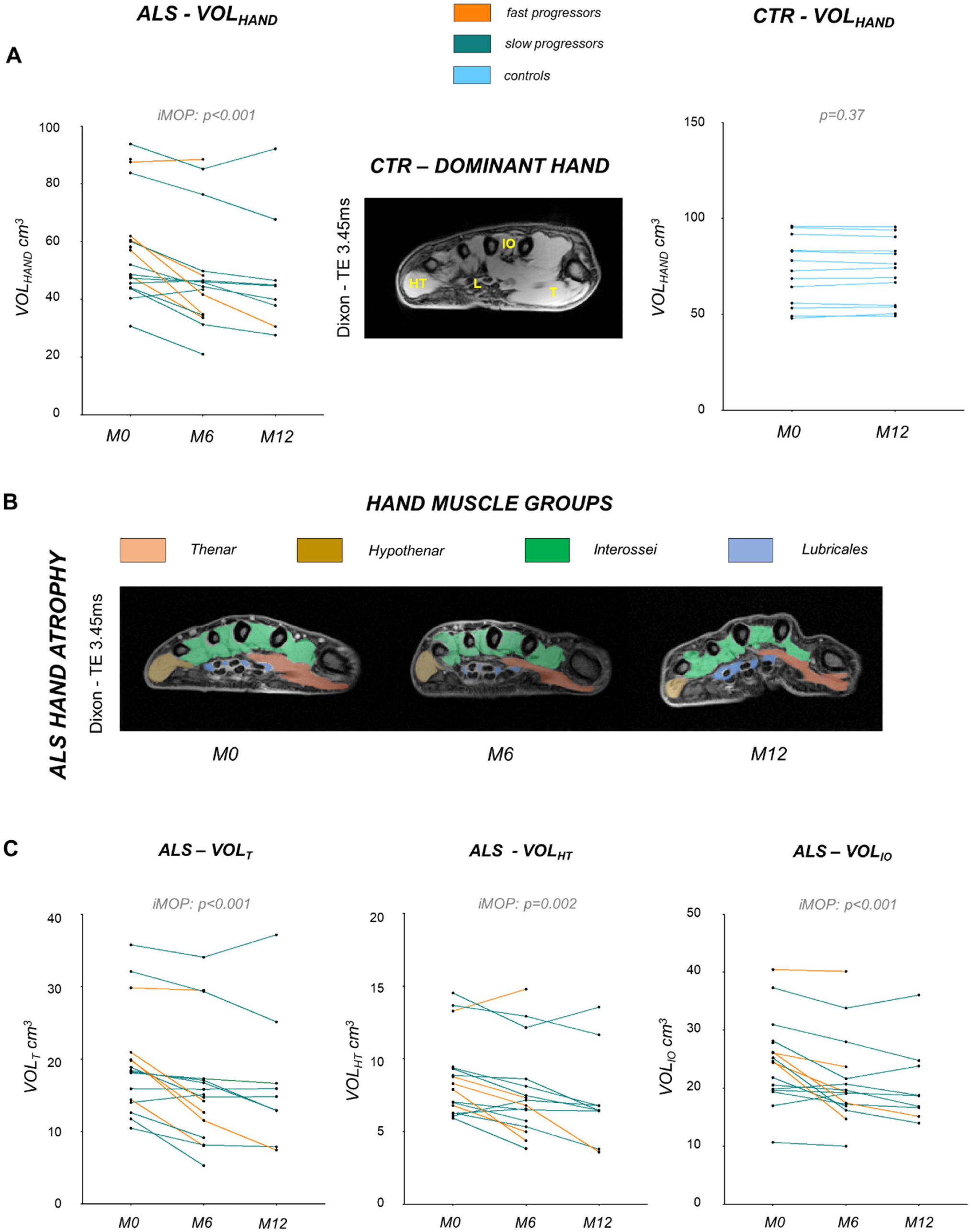
Hand muscle volume detects ALS disease progression. **(A)** Hand volume (VOL*_HAND_*) of ALS patients (left panel; fast progressors: orange; slow progressors: green) and healthy controls (right panel) over time. Data are shown as individual values at baseline (M0), 6 months (M6) and 12 months (M12). VOL*_HAND_* significantly declined over the individual maximum observation period (iMOP) in ALS patients (test statistic=-122, p<0.001; left), while no significant changes were observed in controls (T=0.94, p=0.37, right). Middle panel shows a sample axial Dixon imaging (TE=3.45ms) of the dominant hand of a control. **(B)** Sample of hand muscle segmentation and atrophy on Dixon imaging (TE=3.45ms) at baseline (M0, left), 6 months (M6, middle) and 12 months (M12, right) of the dominant hand of an ALS patient. **(C)** Hand muscle subgroup volumetrics over time in thenar (VOL*_T_*, left), hypothenar (VOL*_HT_*, middle) and interossei muscles (VOL*_IO_*, right). Data are shown as before-after plots of individual values at baseline (M0), 6 months (M6) and 12 months (M12). Volumes of all investigated hand muscles significantly decreased over time (VOL*_T_*: test statistic=-120, p<0.001; VOL*_HT_*: test statistic=-114, p=0.002; VOL*_IO_*: test statistic=-124, p<0.001). Note: ALS: amyotrophic lateral sclerosis; iMOP: individual maximum observation period); M0: baseline; M6: 6-month follow up; M12: 12-month follow-up; VOL*_IO_*: and interossei muscle volume; VOL*_HAND_*: hand volume; VOL*_T_*, left), VOL*_HT_*: hypothenar muscle volume. Muscle abbreviations: HT: hypothenar; IO: interossei; L: lubricales; T: thenar

At head-neck level, bilateral volumetry was performed in the pterygoideus lateralis muscle, revealing a significant decline in muscle volume VOL*_PL_* over the iMOP (partial η^²^=0.47; T(1,15)=3.67, p=0.002), with controls remaining unchanged (**Table 1**, **Figure 2D**).

Highest responsiveness of atrophy measurements was observed for VOL*_HAND_* (SRM 1.17), followed by CSA*_THIGH_* (SRM 1.09) and CSA*_CALF_* (SRM 1.08; **Supplementary Table 3**). We additionally performed supplementary analyses looking only at the interval from baseline to 6 months in all ALS patients and found that even in this time period a decline could be detected in CSA*_THIGH_*, CSA*_CALF_*, VOL*_HAND_*, and VOL*_PL_* (**Figure 1**, **Figure 2**, **Supplementary Figure 1**). Details of these analyses can be found in **Supplementary Table 4**.

### Quantitative muscle MRI parameter fat fraction increases over time

ALS is a fast progressive disease, and we had previously observed a very modest increase in FF in a cross-sectional study^13^. Here, we observed a significant increase in overall FF*_THIGH_* over the iMOP (rs=0.66; test statistic=89.0, p=0.04; **Table 1**, **Figure 3A**) and FF*_CALF_* (rs=0.84; test statistic=123, p=0.002; Fehler! Verweisquelle konnte nicht gefunden werden., **Figure 3B**). Functional rest muscle area (fRMA) significantly decreased both at thigh (fRMA*_THIGH_*; partial η^²^=0.57; T(1,16)=4.57, p<0.001; **Table 1**) and calf level (fRMA*_CALF_*; partial η^²^=0.57; T(1,15)=4.42, p<0.001; **Table 1**). Similarly, a significant increase in overall FF*_HAND_* was observed over iMOP (rs=0.46; test statistic=105.0, p<0.001, **Table 1**). At head-neck level, no significant change in FF*_TONGUE_* was observed over iMOP (partial η^²^=0.11; T(1,13)=1.22, p=0.25; **Table 1**).

**Figure 3.**
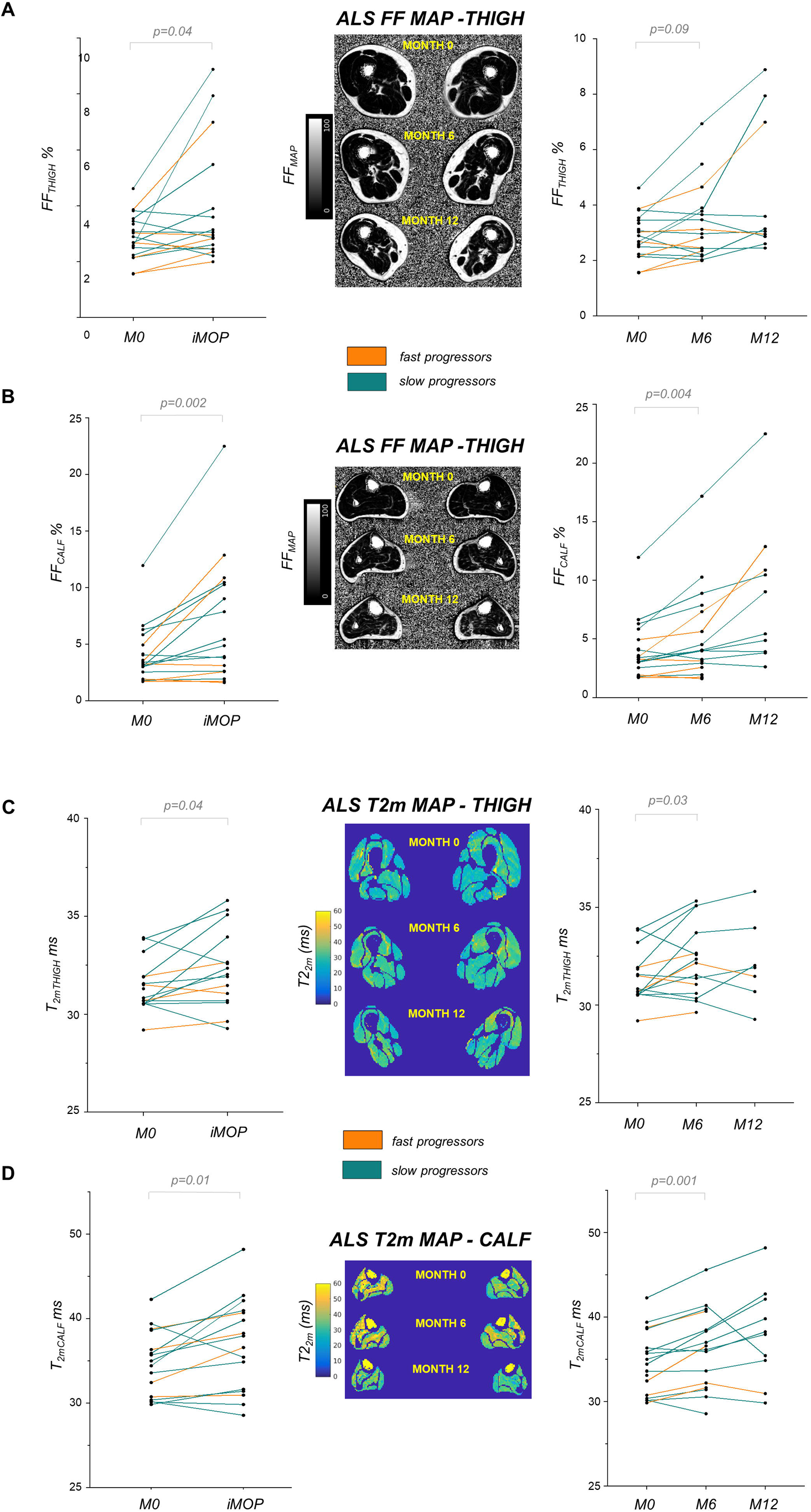
Fat fraction shows increase in ALS thigh and calf muscles. **(A)** Overall muscle fat fraction at thigh level (FF*_THIGH_*; left panel) over time in ALS patients (fast progressors: orange; slow progressors: green). Left panel: mean values at baseline (M0) and individual maximum observation period (iMOP). Right panel: individual values at baseline (M0), 6 months (M6) and 12 months (M12). FF*_THIGH_* significantly increased over the iMOP in ALS patients (test statistic=89.0, p=0.04). At 6-month follow-up, only a trend was observed (test statistic=112.0, p=0.09). Middle panel: Sample FF map axial images of thighs of an ALS patient at baseline (upper row), 6 months (middle row) and 12 months (lower row). **(B)** Overall muscle fat fraction at calf level (FF*_CALF_*) over time in ALS patients. Left panel: mean values at baseline (M0) and individual maximum observation period (iMOP). Right panel: individual values at baseline (M0), 6 months (M6) and 12 months (M12). A significant increase in overall FF*_CALF_* over the iMOP (test statistic=123, p=0.002) was observed. This effect was already significant at 6-month follow-up (test statistic=138.0, p=0.004). Middle panel: Sample FF map axial images of calves of an ALS patient at baseline (upper row), 6 months (middle row) and 12 months (lower row). **(C)** Overall water T2 at thigh level (T_2m_*_THIGH_*) over time in ALS patients. Left panel: mean values at baseline (M0) and individual maximum observation period (iMOP). Right panel: individual values at baseline (M0), 6 months (M6) and 12 months (M12). A significant increase in T_2m_*_THIGH_* over the iMOP (T=-2.33, p=0.04) was observed. This effect was already significant at 6-month follow-up (=-2.37, p=0.03). Middle panel: Sample T2 axial images of thighs of an ALS patient at baseline, 6 months and 12 months. **(D)** Overall water T2 at calf level (T_2m_*_CALF_*) over time in ALS patients. Left panel: mean values at baseline (M0) and individual maximum observation period (iMOP). Right panel: individual values at baseline (M0), 6 months (M6) and 12 months (M12). A significant increase in T_2m_*_CALF_* over the iMOP (T=-2.95, p=0.01) was observed. This effect was already significant at 6-month follow-up (=-4.06, p=0.001). Middle panel: Sample T2 axial images of calves of an ALS patient at baseline, 6 months and 12 months. Note: ALS: amyotrophic lateral sclerosis; FF: fat fraction; FF*_THIGH_*: overall fat fraction of thigh muscles. FF*_CALF_*: overall fat fraction of calf muscles. FF*_ANT-TMC_*: muscle specific fat fraction of anterior thigh muscle compartment; FF*_POSD-CMC_*: muscle specific fat fraction of posterior deep calf muscle compartment; iMOP,:(individual maximum observation period); M0: baseline; M6: 6-month follow up; M12: 12-month follow-up; T_2m_*_CALF_*: overall water T2 at calf level; T_2m_*_THIGH_*: overall water T2 at thigh level.

Highest responsiveness was observed for fRMA*_THIGH_* (SRM 1.11) and fRMA*_CALF_* (SRM 1.10; **Supplementary Table 3**). Supplementary analyses investigating only the 6-month interval from baseline in all ALS patients confirmed changes can be detected in this time frame, with an additional significant effect for FF*_TONGUE_* (**Supplementary Table 4**). In healthy subjects, no changes in overall FF*_CTR_* were observed at thigh, calf, head-neck, or hand level over time (**Table 1**).

### T2 relaxation time as a marker of muscle oedema increases over time

T_2m_ can be used to assess the extent of muscle oedema^19^ and was assessed at calf and thigh level. At baseline, T_2m_ was significantly higher in ALS patients compared to controls both at calf (partial η^²^=0.44, T=5.67, p<0.001) and thigh level (partial η^²^=0.41, T=4.50, p<0.001). In ALS patients, T_2m_ significantly increased over the iMOP at both calf (partial η^²^=0.37, T=-2.95, p=0.01) and thigh level (partial η²=0.28, T=-2.33, p=0.04; **Table 1**, **Figure 3C-D**). Effects were already significant after 6 months (**Supplementary Table 3**). Controls did not show significant changes over time (**Table 1**). The relative increase of T_2m*CALF*_ significantly correlated with the relative increase of FF*CALF* (r=0.70, p=0.003; **Supplementary Figure 2**) over iMOP, while there was no correlation at thigh level (r=0.43, p=0.11; **Supplementary Figure 2**), without significant difference (z=-0.36, p=0.72) between fast progressors (r=0.77, p=0.07) and slow progressors (r=0.65, p=0.04). In controls, no significant changes in T_2m_ were observed over time (**Table 1**).

### Progressive loss of fRMA and muscle volume correlate with disease severity

To assess whether muscle MRI changes are linked and can be used to monitor ALS progression, we correlated relative changes of quantitative MRI measures with muscle strength assessments. We observed a significant correlation of the relative decrease of fRMA*_ANT-TMC_* of the anterior thigh muscle compartment with the relative decrease of knee extension strength over time (r=0.77, p<0.001; **Supplementary Table 5**, **Figure 4A**), without significant difference (z=0.04, p=0.97) between fast progressors (r=0.77, p=0.07) and slow progressors (r=0.76, p=0.01). There was no correlation of the relative change of T_2m_*_THIGH_* with knee extension strength change over time (**Supplementary Table 5**). In the calf, the relative decrease of fRMA*_TRICEPSSUR_* significantly correlated with the relative decrease of plantar flexion strength over the iMOP (r=0.78, p<0.001; **Supplementary Table 5**, **Figure 4B**). Again, there was no difference between fast progressors (r=0.86, p=0.06) and slow progressors (r=0.86, p<0.001; z=-0.03, p=0.98). Furthermore, the relative increase in T_2m_*_CALF_* significantly correlated with the decrease of plantar flexion strength over time (r=0.57, p=0.03, **Supplementary Table 5**, **Supplementary Figure 3**), without differences between fast (r=0.85, p=0.07) and slow progressors (r=0.30, p=0.41; z=.1.37, p=0.17; **Supplementary Table 5**).

**Figure 4.**
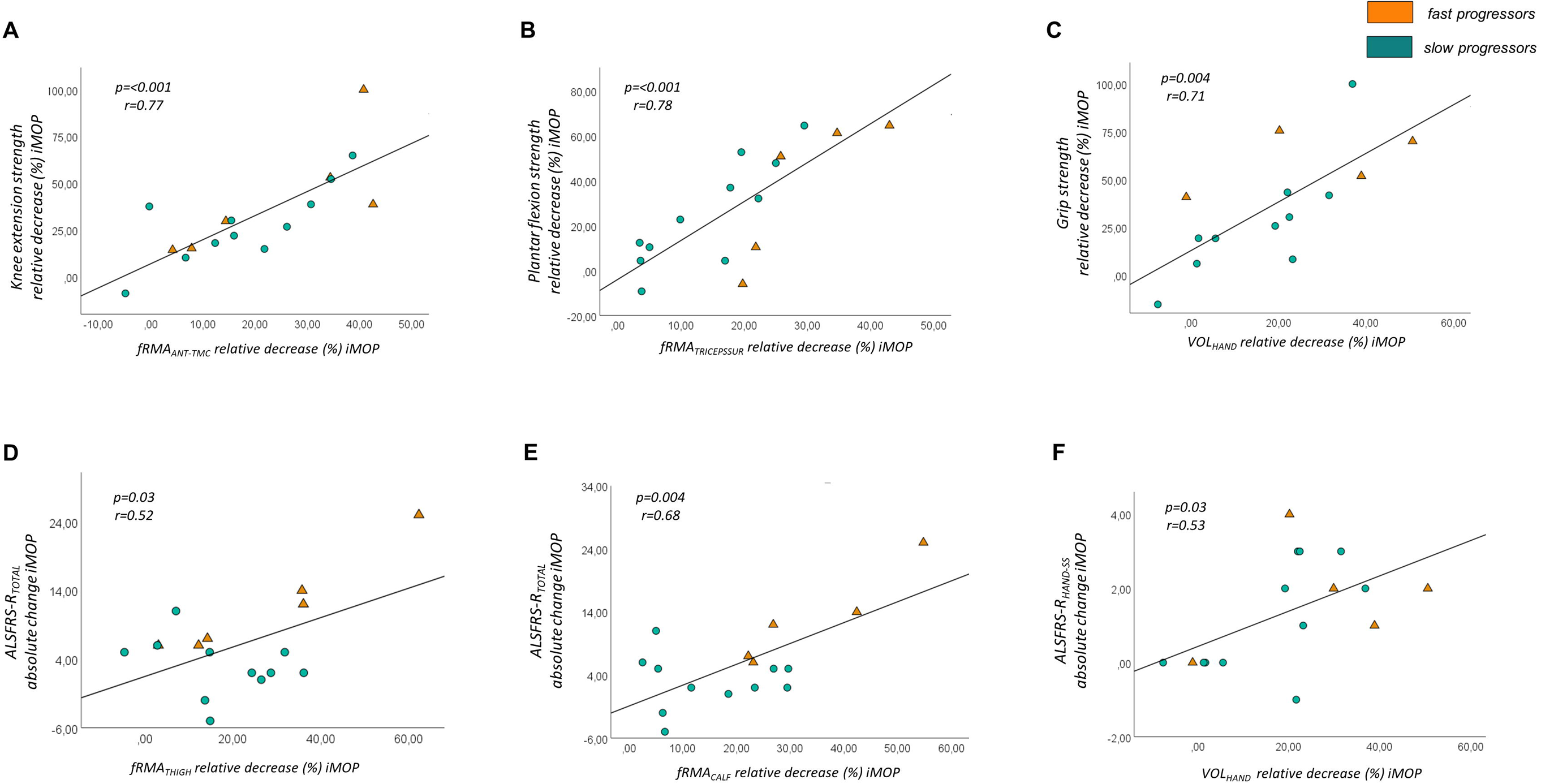
Muscle MRI changes correlate with clinical disease progression in ALS patients. **(A)** At thigh level, a correlation of the relative decrease of fRMA*_ANT-TMC_* with the relative decrease of knee extension strength over the individual maximum observation period (iMOP; r=0.77, p<0.001) was observed, without difference (z=0.04, p=0.97) between fast and slow progressors. **(B)** At calf level, the relative decrease of fRMA*_TRICEPSSUR_* significantly correlated with the corresponding relative decrease of plantar flexion strength over iMOP (r=0.78, p<0.001), without difference (z=-0.03, p=0.98) between fast and slow progressors. **(C)** At hand level, significant correlations were observed between the relative decrease in VOL*_HAND_* and the relative decrease of the grip strength over iMOP (r=0.71, p=0.004), without differences between fast and slow progressors (z=-0.65, p=0.52. **(D)** The relative change of *fRMA_THIGH_* correlated significantly with the absolute change of ALSFRS-R*_TOTAL_* score over the iMOP (r=0.52, p=0.03), with stronger correlations in fast compared to slow progressors (p<0.001, z=3.55). **(E)** The relative change of fRMA*_CALF_* showed a significant correlation with the absolute change of ALSFRS-R_TOTAL_ score over the iMOP (r=0.68, p=0.004), with stronger correlations in fast compared to slow progressors (p=0.01, z=2.70). **(F)** The relative change of VOL*_HAND_* correlated significantly with the absolute change of ALSFRS-R*_HAND-SS_* subscore over iMOP (r=0.53, p=0.03), without differences between fast and slow progressors (z=-0.61, p=0.27). Note: ALS: amyotrophic lateral sclerosis; ALSFRS-R: ALS functional rating scale – revised; ALSFRS-R*_TOTAL_*: ALS functional rating scale total score; ALSFRS-R*_HAND-SS_*: ALS functional rating scale hand subscale score; fRMA*_THIGH_*: overall functional remaining muscle area of thigh muscles; fRMA*_CALF_*: overall functional remaining muscle area of calf muscles; fRMA*_ANT-TMC_*: muscle specific functional remaining muscle area of anterior thigh muscle compartment; fRMA*_TRICEPSSUR_*: muscle specific functional remaining muscle area of triceps surae muscles; VOL*_HAND_*: overall volume of the hand muscles; iMOP, individual maximum observation period.

We then assessed whether MRI findings correlated with functional rating scales and observed a significant correlation between the overall relative decrease of fRMA and decrease of the ALSFRS*_TOTAL_* score over the iMOP at both thigh level (r=0.52, p=0.03; **Supplementary Table 5**, **Figure 4C**) and calf level (r=0.68, p=0.004), **Supplementary Table 5**, **Figure 4D**). Correlations were stronger in fast progressors (r=0.97, p=0.001) than slow progressors (r=-0.30, p=0.37) at thigh level (p<0.001, z=3.55), and in fast progressors (r=0.95, p=0.01) than slow progressors (r=0.001, p=0.99) at calf level (p=0.01, z=2.70). Correlations with ALSFRS*_LL-SS_* and fRMA of thighs and calves remained non-significant (thigh: r=0.39, p=0.12; calf: r=0.16, p=0.55, **Supplementary Table 5**), although we noticed a significant correlation in subanalyses of fast progressors at thigh level (r=0.87, p=0.02, **Supplementary Table 5**). Interestingly, we observed a correlation of T_2m_*_THIGH_* and functional rating scale scores (ALSFRS*_TOTAL_* and ALSFRS*_LL-SS_*), indicating an increase in T_2m_*_THIGH_* relating to higher scores on functional rating scales (**Supplementary Table 5**, **Supplementary Figure 3**), without differences between fast and slow progressors (ALSFRS*_TOTAL_* z=0.63, p=0.59, ALSFRS*_LL-SS_* z=0.22, p=0.83). At calf level, there was no significant correlation of relative changes in T_2m_*_CALF_* with ALSFRS*_TOTAL_* or ALSFRS*_LL-SS_* scores (**Supplementary Table 5**).

We investigated the relation between VOL*_HAND_* changes and hand grip strength. The relative decrease of VOL*_HAND_* significantly correlated with the corresponding relative decrease of grip strength over time (r=0.71, p=0.004, **Supplementary Table 5**, **Figure 4E**), without differences between fast (r=0.52, p=0.48) and slow progressors (r=0.80, p=0.005; z=-0.65, p=0.52, **Supplementary Table 5**). We also tested for correlations between VOL*_HAND_* and ALSFRS*_HAND-SS_*; revealing a correlation between these parameters over time (r=0.53, p=0.03, **Supplementary Table 5**, **Figure 4F**), without differences between fast progressors (r=0.27, p=0.66) and slow progressors (r=0.64, p=0.03; z=-0.61, p=0.27, **Supplementary Table 5**). Also at head-neck level, longitudinal MRI changes correlated with changes in ALSFRS*_BULBAR-SS_* over time (Supplementary material).

## DISCUSSION

We have evaluated the potential of muscle MRI as an imaging biomarker of disease progression in ALS. By applying quantitative muscle MRI measures of atrophy, oedema and fat infiltration, we observed significant changes in all assessed regions over time. Furthermore, we found a correlation between changes in quantitative MRI parameters and clinical disease severity, in both fast and slow progressors. In general, we observed a significant progressive atrophy of the dominant hand, lower limbs and bulbar muscles over time. Additionally, we noticed an increase in muscle fat accumulation of the lower limbs, with the most pronounced changes in the distal leg muscles.

Previous longitudinal investigations of muscle MRI in ALS are few, and mostly limited to qualitative MRI parameter changes^20^, small sample sizes^21^, or changes in relative T2 signal over short timescales^22^, offering alternative diagnostic methods for muscles inaccessible to electrophysiology^19^. However, previous studies failed to demonstrate the quantification of progressive muscle atrophy in large patient groups^23^, or did not report any longitudinal MRI parameter changes, along with a lack of correlation with clinical disease severity^24^. In this study, we provide reproducible and previously validated^13,17^ quantitative muscle MRI protocols for multiple anatomic regions, which is particularly important, given the diverse patterns of anatomical onset and highly variable disease course of ALS.

Since progressive muscle atrophy is one of the main clinical features of ALS, the quantification of muscle wasting is of upmost importance to objectively monitor disease progression. In a pilot study, Jenkins et al. proposed volumetric muscle MRI as a potential biomarker to monitor disease progression in ALS^21^. One of the first muscle MRI studies in ALS observed abnormalities in size, shape and position of the tongue, without comparison to healthy controls^25^. Two decades later, a cross-sectional study found evident atrophy, and muscle fat infiltration in ALS was assessed on conventional MRI sequences such as T1-weighted MR imaging, along with alterations in T2-weighted and short-tau inversion recovery (STIR) signal intensities in subscapularis and supra/infraspinatus muscles^26^. By applying T1-weighted imaging, Diamanti et al.^27^ reported significant atrophy of the hands of ALS patients, compared to healthy controls.

For the first time, we applied longitudinal quantitative MRI measures of muscle atrophy (CSA and muscle volume) at multiple anatomical regions and detected progressive, quantifiable muscle wasting in ALS patients. Progressive muscle atrophy was evident in all investigated regions, including head and neck muscles, but was most marked in the dominant hand (SRM -1.17) and leg muscles (SRM thigh: -1.09, SRM calf: -1.08). These findings are supported by recently published longitudinal data on muscle fat fraction in ALS suggesting a primary involvement of calf muscles^28^. We extend on these findings by quantification of progressive muscle atrophy and additionally analysing hand and head-neck-level muscles as well as correlating MRI changes with both functional rating scales and muscle strength measurements.

T_2m_ increased over time both at thigh and calf level as an expression of fluid shifts caused by active muscle denervation^29^. At calf level, the progressive increase in T_2m_ correlated with the progressive loss of clinical muscle strength, which is in line with a previous longitudinal investigation using a whole-body MRI approach^22^. At thigh level, on the contrary, we observed a correlation between higher T_2m_ and functional rating scale scores. While this might seem counterintuitive, similar effects have recently been reported by Paoletti et al.^28^, where this interrelation was attributed to the course of disease progression, indicating that function may be preserved in earlier phases of the disease (particularly in proximal leg muscles), where active denervation and increase of T_2m_ is predominant, while T_2m_ may show progressive decrease in later disease stages. We therefore argue that our findings at thigh level reflect earlier stages of the disease, where clinical impairment is not as prominent, while calf muscles reflect a more advanced stage of disease, where progressive muscle oedema leads to functional impairment evidenced by loss of muscle strength, which is also in line with previous observations suggesting a primary involvement of calf muscles in ALS^22,28^.

Due to the natural course of this progressive and fatal disease, data from approximately half of the initially enrolled patients were not available at the end of follow-up. Furthermore, patients available for follow-up scans already had difficulties with the ability to lie still during MRI acquisitions, considering the extended MRI protocols to image multiple anatomical regions. Currently, muscle MRI could be of use in ALS populations in early disease stages who are more likely to tolerate scanning over a six-month period. Quantifying muscle wasting by applying conventional T1-weighted sequences can reduce the costs and duration of scanning time, which is important considering the inability of ALS patients to maintain the desirable position for long periods. Further work will be necessary to implement shorter scanning protocols and upright positioning in order to be applicable to subsequent disease stages.

Recently, Jenkins et al.^22^ reported the first whole-body MRI application in ALS, with T2-weighted imaging of distal legs being more sensitive than diffusion-weighted sequences in detecting longitudinal group-level changes. Volumetric muscle MRI, particularly the examination of progressive atrophy of the dominant hand, appears to provide comparable features of an imaging biomarker of disease progression in all ALS patients, irrespective of the onset of anatomical region, with the advantage of much shorter scanning times compared to whole-body MRI. As we observed significant quantitative muscle MRI parameter changes already at 6-month follow-up, future study designs could include shorter intervals between scans, along with investigations at higher field strengths (7 Tesla).

By applying quantitative MRI to multiple anatomical regions, we demonstrate the full potential of muscle MRI to reflect the disease severity in MND as we were able to show that changes in MRI obtained parameters correlate strongly with established clinical measures of disease progression. We also provide novel MRI protocols for assessing progressive atrophy of the hand and bulbar region, both crucial in tracking the disease progression in such conditions. Biomarkers as urinary p75^ECD^ have also been useful in determining disease progression^30^, but do not have the spatial resolution offered by MRI – it is likely that complementing these approaches will be beneficial.

Patient survival is frequently used as an endpoint in clinical trials of ALS^31^, but objective markers of disease progression may reduce the size and duration of future trials. MRI-quantified atrophy of the hand and lower limbs showed the highest responsiveness to detect disease progression, further validated by strong correlation with patient muscle strength and functional scores. Our findings might have substantial impact on the design of future clinical trials in ALS, potentially allowing a reduced number of participants needed to detect effects, with the ultimate goal of facilitating the development and approval of therapeutic agents in ALS. There is an urgent need for the development of new drugs in ALS and our longitudinal study contributes to a small but fast rising field of muscle MRI in MND and supports its integration in future interventional trials.

## Data Availability

Data available: No.
Additional Information: Explanation for why data not available: The data that support the findings of this study are controlled by the respective centers and are not publicly available. Request to access the raw data should be forwarded to data controllers via the corresponding author. Written requests for access to the derived data will be considered by the corresponding author and a decision made about the appropriateness of the use of the data.

## ACKNOWLEDGEMENTS

We thank all study participants and families for their participation to research that has made this work possible. This study was supported by the UCLH NIHR Biomedical Research Centre. UK is funded by KD-UK. PF is funded by an MRC and MNDA LEW Fellowship. LG is the Graham Watts Senior Research Fellow funded by the Brain Research Trust.

## DATA SHARING STATEMENT

Data available: No.

Additional Information: Explanation for why data not available: The data that support the findings of this study are controlled by the respective centers and are not publicly available. Request to access the raw data should be forwarded to data controllers via the corresponding author. Written requests for access to the derived data will be considered by the corresponding author and a decision made about the appropriateness of the use of the data.

## AUTHOR CONTRIBUTIONS

Dr Uros Klickovic had full access to all of the data in the study and takes responsibility for the integrity of the data and the accuracy of the data analysis.

Concept and design: Klickovic, Morrow, Thornton, Fratta

Acquisition, analysis, or interpretation of data: All authors.

Drafting of the manuscript: Klickovic, Fratta

Critical review of the manuscript for important intellectual content: All authors

Statistical analysis: Klickovic, Trimmel

Obtained funding: Fratta

Administrative, technical, or material support: Zampedri

Supervision: Morrow, Thornton, Fratta

Other: Orrell

