## Supplementary for "Muscle MRI quantifies disease progression in Amyotrophic Lateral Sclerosis"

### Supplementary Material

#### Methods

***Study Design and patient recruitment***

We performed a prospective longitudinal observational study in 20 consecutive male and female patients with ALS who were recruited from the motor neuron disease clinic at the National Hospital for Neurology and Neurosurgery, Queen Square, London, UK between 2015 and 2017. All patients had a history of at least clinically probable disease according to revised El Escorial criteria[1] (12 probable, 8 definitive). Out of 20 ALS patients, 17 patients were available for 6-month follow-up and 10 patients for 12-month follow-up, which represented the individual maximum observation period (iMOP). The cohort was furthermore stratified according to the monthly decrease in ALS Functional Rating Scale - Revised (ALSFRS-R) score into slow (≤0.5 point per month, n=11), and fast (≥ 1.0 point per month, n=6) disease progressors. Sixteen healthy controls with matched demographic characteristics formed the control group. Exclusion criteria were concomitant neuromuscular diseases and MRI safety-related contraindications. ALS patients received clinical assessments and muscle MRI of the hand, thighs, calves, and head-neck region at baseline as well as 6-month and 12-month follow-up. Healthy controls were examined at baseline and at 12-month follow-up (iMOP). Cross-sectional data of baseline measurements in ALS patients and healthy controls, combined with data from other study participants, have been reported previously[2].

At baseline, thigh $T_{2m}$ data of 2 patients and both thigh and calf $T_{2m}$ data of 1 had to be excluded due to poor image quality. For baseline FF hand analyses, data of 3 patients (of which one was not available for follow-up) and 3 controls was excluded due to technical issues with image processing, and 1 patient because of incomplete scan examination. Four ALS patients were excluded from hand correlation analyses as they were unable to perform myometry assessments at baseline and floor effects prevented identification of a longitudinal decline. At 6-month follow-up, thigh and calf $T_{2m}$ data of 1 patient had to be excluded due to poor image quality and hand imaging could not be acquired in 1 patient due to inability to lie still in the scanner. At head-neck level, 2 patients were excluded from tongue FF analyses, of which 1 also had to be excluded from muscle volumetrics and 1 patient from FF analyses only due to poor image quality from excessive movement artifacts. At 12-month follow-up, thigh and calf $T_{2m}$ data of 2 controls were excluded to poor image quality and head-neck imaging of 2 further patients and one control was excluded because of artifacts. FF hand analyses of one control had to be excluded due to incomplete scan examination. Hand and head-neck imaging of one additional patient could not be acquired due to inability to lie still in the scanner.

Out of 16 healthy controls, 15 were available for follow-up. At follow-up, hand imaging of two controls was excluded, with one due to insufficient image quality, and another due to incomplete scan examination.

#### *Standard Protocol Approvals, Registrations, and Patient Consents*

The UCL Research Ethics Committee gave ethical approval for this work (11/LO/1425) and written informed consent was obtained from all participants.

#### *Data acquisition*

#### *Clinical and functional testing*

Patients and healthy controls were functionally rated using the ALS Functional Rating Scale-Revised (ALSFRS-R)[3]. The ALSFRS-R consists of 12 questions that each have a score from 0-4, where 4 indicates normal function and 0 indicates no function[3]. All study participants underwent detailed upper and lower limb myometry on a wireless handheld microFET®2 handheld dynamometer (Hoggan Scientific; UT, USA) for isometric assessment of jaw opening, wrist extension and flexion, finger extension and flexion, thumb abduction and adduction, knee extension and flexion, ankle extension and flexion. Handgrip strength and lateral pinch (between the radial side of the index finger and the thumb) were assessed with a Martin Vigorimeter (GP Supplies; London, UK)[4]. Except for jaw opening, all measurements were performed bilaterally. Iowa-Oral-Pressure-Instrument (IOPI) measures were carried out to asses the tongue pressure and bilateral peri-oral pressure[5]. IOPI measures tongue strength by using an inflatable tongue bulb with a pressure sensor. All asessments consisted of 3 attempts of 3-5 seconds of which the best attempt was selected. IOPI could not be acquired in four patients at baseline due to technical issues. All patients and controls additionally received detailed clinical assessments including medical history as well as clinical and neurological examinations.

#### *MR Imaging*

Images of the participants' thighs, calves, hand and head-neck region were acquired at on a 3 Tesla Skyra MR system (Siemens Healthineers, Erlangen, Germany). Quantitative fat-fraction maps of the lower limbs, hand and head-neck were produced from the 3-point Dixon images (Thighs and calves: 2D gradient echo, nine 6 mm slices, TR = 102 ms, TE = 3.45/4.60/5.75 ms, flip-angle 10°, NSA = 4, FOV = 420 x 180 mm, voxel size 1.3 x 1.3 x 6 $mm^3$; Hand: 2D gradient echo, nine 6 mm slices, TR = 102 ms, TE = 3.45/4.60/5.75 ms, flip-angle 10°, NSA = 4, FOV = 180 x 90 mm, voxel size 0.56 x 0.56 x 6 $mm^3$; Head-neck: 2D gradient echo, eleven 10 mm slices, TR = 125 ms, TE = 3.45/4.60/5.75 ms, flip-angle 10°, NSA = 3, FOV = 235 x 180 mm, voxel size 0.56 x 0.56 x 10 $mm^3$) as described in detail previously[2]. T2 relaxometry data were acquired with a multi-echo-spin-echo sequence (Thighs and calves: Nine 6 mm slices, TR = 3500 ms, 22 echoes TE = 10 to 220 ms in 10 ms steps, NSA = 1 FOV = 420 x 180 mm, voxel size 1.3 x 1.3 x 6 $mm^3$).

Total scan time for all sequences at multiple anatomical regions, including patient re-positioning, was about 60 minutes.

#### *MRI Data Analysis*

A single experienced radiologist (U.K.) blinded to study groups outlined the muscles in the hand, thighs, calves and head-neck region using ITK-SNAP[6]. These regions were used to calculate quantitative MRI measures fat fraction (FF), cross-sectional area (CSA), and functional remaining muscle area (fRMA) for each muscle and muscle compartment group as well as a weighted overall mean muscle FF, described in detail previously[2]. WaterT2 ($T_{2m}$) estimation: A multi-component, slice profile-corrected EPG model [s(TE) = (1 - ffa) · sEPG(B1f, $T_{2m}$, α, σN, TE) + ffa · [ 0.33 · sEPG(B1f, T2=40ms, α, σN, TE) + 0.67 · sEPG(B1f, T2=198ms, α, σN, TE)] was fitted pixel-wise to the data using maximum likelihood estimation in MATLAB, to estimate $T_{2m}$ and an apparent fat fraction ffa. The 2-component fat-signal model parameters were estimated a priori from 4 subcutaneous fat ROIs in 8 representative subjects. Muscles were manually segmented on single slices at thigh and calf level. Mean $T_{2m}$ and ffa were calculated for the entire musculature cross section at each level. Additionally, a manual slice-by-slice segmentation was performed on 2D gradient echo Dixon sequences (TE 3.45ms) to calculate the muscle volume of the dominant hand.

#### *Statistical Analysis*

Statistical analyses were performed with GraphPad Prism version 9.1.2 with a p-value threshold of 0.05. As appropriate to the distribution of data, measures are reported as mean ± SD or median ± interquartile range (IQR). For between-group comparisons, 2-sample t-tests or Mann-Whitney-U-tests were applied, and for within-group comparisons (change over time in individual maximum observation period), paired t-tests or Wilcoxon signed-rank tests were applied as appropriate. Missing data were excluded from analyses. Correlations of MRI data with clinical measures were investigated with Spearman (rho) or Pearson coefficients as appropriate for the distribution of data. Differences between correlations were assessed with Fisher`s r to z transformation. Effect sizes are reported as partial $\eta^2$ respectively Spearman's Rank Correlation Coefficient. MRI data responsiveness was assessed using standardized response mean (SRM; mean change divided by the change SD).

**Supplementary Table 1.** Demographic data in ALS patients and controls. Data are presented as mean±SD or median (IQR) according to data distribution.

| | **ALS (n=20)** | **Controls (n=16)** | **p** |
|---|---|---|---|
| Sex (male/female) | 14/6 | 11/5 | 0.61 |
| Handedness (right/left) | 19/1 | 15/1 | 0.70 |
| Age (years) | 60.5 (24.5) | 62.0 (24.5) | 0.72 |
| BMI | 22.9±3.1 | 26.4±4.0 | **0.01** |
| Disease duration (years) | 2.0 (1.4) | n.a. | n.a. |
| Bulbar involvement (yes/no) | 8/12 | n.a. | n.a. |
| Riluzole therapy (yes/no) | 15/5 | n.a. | n.a. |

**Supplementary Table 2.** Functional rating scales and myometry results in ALS patients at baseline and follow-up (individual maximum observation period, iMOP). Data are presented as mean±SD or median (IQR) according to data distribution.

| | **Baseline** | **Follow-up** | **p** |
|---|---|---|---|
| ALSFRS-$R_{TOTAL}$ | 41.0 (2.0) | 35.0 (10.0) | **<0.001** |
| ALSFRS-$R_{LL\text{-}SS}$ | 6.0 (3.5) | 4.0 (3.0) | **0.01** |
| ALSFRS-$R_{BULBAR\text{-}SS}$ | 12.0 (2.5) | 11.0 (4.5) | **0.01** |
| ALSFRS-$R_{HAND\text{-}SS}$ | 6.0 (1.5) | 4.0 (4.0) | **0.002** |
| Knee extension (Nm) | 59.5±18.3 | 36.4±17.4 | **<0.001** |
| Knee flexion (Nm) | 59.8±22.7 | 32.7±18.7 | **<0.001** |
| Dorsal extension (Nm) | 36.4±21.5 | 23.3±19.7 | **<0.001** |
| Plantar flexion (Nm) | 35.7±14.4 | 24.2±15.9 | **0.001** |
| Hand grip (Nm) | 124.7±45.1 | 76.9±47.9 | **<0.001** |
| Hand pinch (Nm) | 46.5±37.7 | 28.4±31.4 | **<0.001** |
| Jaw opening (Nm) | 17.9±5.1 | 12.1±1.9 | **0.001** |
| IOPI (kPa) | 40.4±14.2 | 35.5±15.8 | 0.13 |

**Supplementary Table 3**. Change in quantitative MRI parameters between baseline vs. 6 months and baseline vs. individual maximum observation period (iMOP) in ALS patients.

| | baseline vs. 6 months | | | baseline vs. iMOP | | |
|---|---|---|---|---|---|---|
| **Atrophy** | **Mean change** | **SD** | **SRM** | **Mean change** | **SD** | **SRM** |
| $CSA_{CALF}$ ($cm^2$) | -15.06 | 17.63 | -0.85 | -22.64 | 20.89 | -1.08 |
| $CSA_{THIGH}$ ($cm^2$) | -24.34 | 24.44 | -1.00 | -38.12 | 34.82 | -1.09 |
| $VOL_{HAND}$ ($cm^3$) | -8.37 | 7.29 | -1.15 | -10.56 | 9.01 | -1.17 |
| $VOL_{T}$ ($cm^3$) | -3.28 | 2.93 | -1.12 | -4.15 | 3.72 | -1.12 |
| $VOL_{HT}$ ($cm^3$) | -1.04 | 1.28 | -0.81 | -1.64 | 1.59 | -1.03 |
| $VOL_{IO}$ ($cm^3$) | -3.42 | 3.67 | -0.93 | -3.98 | 3.97 | -1.00 |
| $VOL_{PL}$ ($cm^3$) | -0.60 | 0.73 | -0.83 | -0.96 | 1.05 | -0.92 |
| **Fat infiltration** | **Mean change** | **SD** | **SRM** | **Mean change** | **SD** | **SRM** |
| $FF_{CALF}$ (%) | 1.30 | 1.71 | 0.76 | 2.75 | 3.38 | 0.81 |
| $FF_{THIGH}$ (%) | 0.47 | 0.82 | 0.58 | 1.02 | 1.73 | 0.59 |
| $FF_{HAND}$ (%) | 1.59 | 2.79 | 0.57 | 1.97 | 3.26 | 0.61 |
| $FF_{TONGUE}$ (%) | 5.93 | 8.03 | 0.74 | 4.15 | 8.90 | 0.47 |
| $fRMA_{CALF}$ ($cm^2$) | -15.61 | 17.59 | -0.89 | -24.08 | 21.80 | -1.10 |
| $fRMA_{THIGH}$ ($cm^2$) | -24.12 | 23.98 | -1.01 | -37.92 | 34.22 | -1.11 |
| $fRMA_{TONGUE}$ ($cm^2$) | -1.88 | 4.71 | -0.40 | -1.05 | 5.68 | -0.19 |
| **$T_{2m}$** | **Mean change** | **SD** | **SRM** | **Mean change** | **SD** | **SRM** |
| $T2m_{CALF}$ (ms) | 1,58 | 1,56 | 1,02 | 2,27 | 3,07 | 0,74 |
| $T2m_{THIGH}$ (ms) | 0,82 | 1,34 | 0,61 | 0,87 | 1,45 | 0,60 |

**Supplementary Table 4**. Quantitative MRI parameters in ALS patients at baseline and 6-month follow-up. Data are presented as mean±SD or median (IQR) according to data distribution.

| **Atrophy** | **Baseline** | **6-month follow-up** | **p** | **95% CI** |
|---|---|---|---|---|
| $CSA_{CALF}$ ($cm^2$) | 112.80±33.17 | 100.69±28.92 | **0.004** | -24.46 – -5.67 |
| $CSA_{THIGH}$ ($cm^2$) | 183.33±59.24 | 161.86±57.92 | **<0.001** | -39.61 – -11.78 |
| $VOL_{HAND}$ ($cm^3$) | 57.04 (16.39) | 45.18 (14.82) | **<0.001** | -12.25 – -1.28 |
| $VOL_{T}$ ($cm^3$) | 18.33 (6.59) | 14.92 (7.48) | **<0.001** | -6.44 – -0.81 |
| $VOL_{HT}$ ($cm^3$) | 8.30 (2.65) | 6.98 (3.06) | **0.01** | -0.85 – -0.25 |
| $VOL_{IO}$ ($cm^3$) | 25.22 (8.67) | 19.26 (6.18) | **0.001** | -6.52 – -0.48 |
| $VOL_{PL}$ ($cm^3$) | 14.24±3.92 | 13.64±3.64 | **0.002** | -0.99 – -0.21 |

| **Fat infiltration** | **Baseline** | **6-month follow-up** | **p** | **95% CI** |
|---|---|---|---|---|
| $FF_{CALF}$ (%) | 3.34 (2.07) | 3.99 (4.84) | **0.004** | 0.42–2.18 |
| $FF_{THIGH}$ (%) | 2.79 (1.13) | 2.96 (1.56) | 0.09 | 0.05–0.90 |
| $FF_{HAND}$ (%) | 2.22 (3.47) | 5.54 (4.10) | 0.07 | -0.18–3.36 |
| $FF_{TONGUE}$ (%) | 8.50± 3.42 | 14.10 (14.11) | **0.02** | 1.08–10.78 |
| $fRMA_{CALF}$ ($cm^2$) | 108.48±32.67 | 95.90±29.31 | **0.003** | 6.24–2.50 |
| $fRMA_{THIGH}$ ($cm^2$) | 178.12±57.59 | 156.81±56.83 | **<0.001** | 11.79–36.45 |
| $fRMA_{TONGUE}$ ($cm^2$) | 25.94±4.97 | 23.47±5.28 | 0.09 | -0.97–4.73 |

| **$T_{2m}$** | **Baseline** | **6-month follow-up** | **p** | **95% CI** |
|---|---|---|---|---|
| $T_{2mCALF}$ (ms) | 34.61±3.85 | 36.19±4.65 | **0.001** | -2.42 – -0.75 |
| $T_{2mTHIGH}$ (ms) | 31.43±1.34 | 32.25±1.85 | **0.03** | -1.56 – -0.08 |

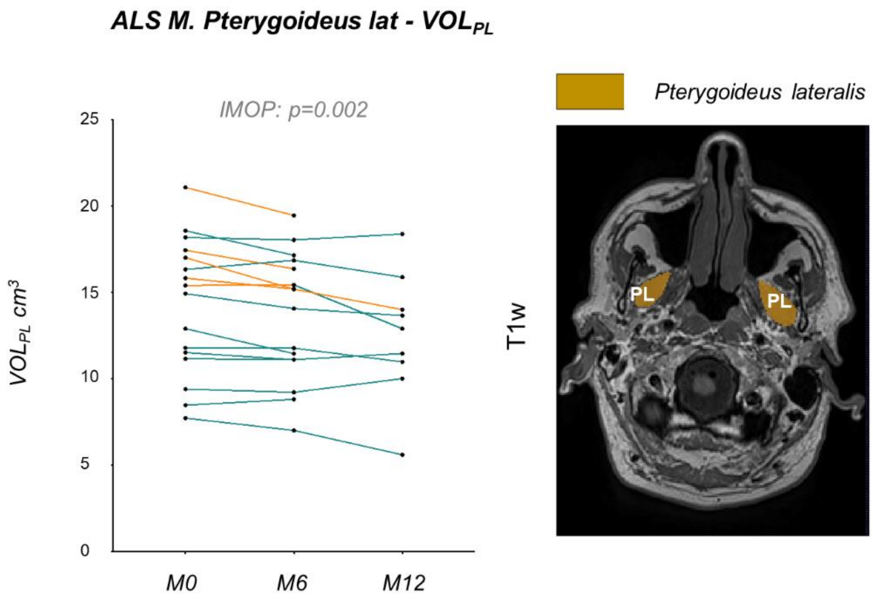


**Supplementary Figure 1.** Volume of the bilateral pterygoideus lateralis muscles (VOL$_{PL}$, left) with corresponding T1w sample image of an ALS patient (right). Data are shown as before-after plots of individual values at baseline (M0), 6 months (M6) and 12 months (M12). A significant decline in muscle volume VOL$_{PL}$ was observed over the individual maximum observation period (iMOP; $T(1,15)=3.66$, $p=0.002$).

Note: VOL$_{PL}$: muscle specific volume of pterygoideus lateralis muscles, iMOP: individual maximum observation period).

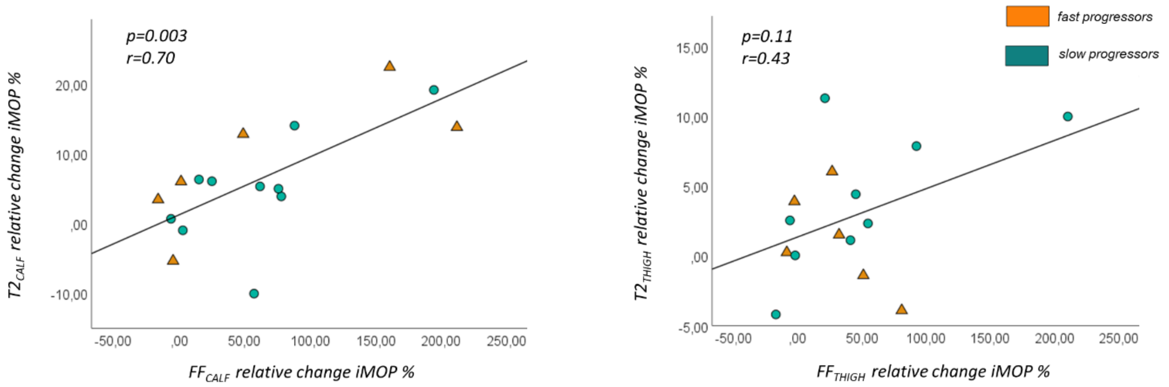


**Supplementary Figure 2**. Correlation of longitudinal water T2 changes vs. muscle fat fraction (FF) changes over individual maximum observation period (iMOP). Left panel: The relative increase of $T_{2mCALF}$ significantly correlated with the relative increase of $FF_{CALF}$ (left panel; r=0.70, p=0.003) over iMOP, without significant difference (z=-0.36, p=0.72) between fast progressors (r=0.77, p=0.07) and slow progressors (r=0.65, p=0.04). Right panel: No significant correlation was observed at thigh level (r=0.43, p=0.11).

Note: FF: muscle fat fraction; $T_{2mCALF:}$ water T2m at calf level; $T_{2mTHIGH}$: water T2 at thigh level.

**Supplementary Table 5**. Correlations of longitudinal changes of quantitative MRI parameters with changes in functional rating scales and muscle strength assessments over time in ALS patients.

| Functional rating scales | Correlation coefficient | p | 95%CI |
|---|---|---|---|
| $ALSFRS_{TOTAL}$ × $fRMA_{THIGH}$ | 0.52 | 0.03 | 0.05 – 0.80 |
| fast progressors | 0.97 | 0.001 | 0.75 – 1.00 |
| slow progressors | -0.30 | 0.37 | 0.76 – 0.37 |
| $ALSFRS_{TOTAL}$ × $fRMA_{CALF}$ | 0.68, | 0.004 | 0.28 – 0.88 |
| fast progressors | 0.95 | 0.01 | 0.43 – 1.00 |
| slow progressors | 0.001 | 0.99 | 0.60 – 0.60 |
| $ALSFRS_{LL-SS}$ × $fRMA_{THIGH}$ | 0.39 | 0.12 | 0.12 – 0.37 |
| fast progressors | 0.87 | 0.02 | 0.19 – 0.99 |
| slow progressors | -0.12 | 0.74 | 0.67 – 0.52 |
| $ALSFRS_{LL-SS}$ × $fRMA_{CALF}$ | 0.16 | 0.55 | -0.36 – 0.61 |

| | | | |
|---|---|---|---|
| fast progressors | -0.16 | 0.79 | 0.91 – 0.84 |
| slow progressors | 0.09 | 0.80 | 0.54 – 0.65 |
| $ALSFRS_{TOTAL}$ × $T_{2mTHIGH}$ | -0.60 | 0.02 | -0.85 – -0.13 |
| fast progressors | -0.62 | 0.19 | -0.95 – 0.39 |
| slow progressors | -0.61 | 0.08 | -0.91 – 0.09 |
| $ALSFRS_{LL-SS}$ × $T_{2mTHIGH}$ | -0.60 | 0.02 | -0.85 – -0.13 |
| fast progressors | -0.48 | 0.34 | -0.93 – 0.55 |
| slow progressors | -0.59 | 0.101 | -0.90 – 0.13 |
| $ALSFRS_{TOTAL}$ × $T_{2mCALF}$ | 0.36 | 0.17 | -0.16 – 0.73 |
| fast progressors | 0.63 | 0.18 | -0.38 – 0.95 |
| slow progressors | -0.16 | 0.65 | -0.06 – 0.88 |
| $ALSFRS_{LL-SS}$ × $T_{2mCALF}$ | 0.28 | 0.30 | -0.14 – 0.74 |
| fast progressors | 0.64 | 0.17 | -0.36 – 0.96 |
| slow progressors | -0.06 | 0.88 | -0.66 – 0.59 |
| $ALSFRS_{HAND-SS}$ × $VOL_{HAND}$ | 0.53 | 0.03 | 0.05 – 0.81 |
| fast progressors | 0.27 | 0.66 | -0.80 – 0.93 |
| slow progressors | 0.64 | 0.03 | 0.07 – 0.90 |
| $ALSFRS_{BULBAR-SS}$ × $fRMA_{TONGUE}$ | 0.78 | 0.003 | 0.38 – 0.93 |
| fast progressors | n.a. | n.a. | n.a. |
| slow progressors | n.a. | n.a. | n.a. |
| $ALSFRS_{BULBAR-SS}$ × $VOL_{PL}$ | 0.45 | 0.04 | -0.07 – 0.78 |
| fast progressors | 0.44 | 0.46 | -0.73 – 0.95 |
| slow progressors | 0.37 | 0.35 | -0.35 – 0.79 |

| **Muscle strength assessments** | **Correlation coefficient** | **p** | **95%CI** |
|---|---|---|---|
| Knee extension × $fRMA_{ANT-TMC}$ | 0.77 | 0.001 | 0.46 – 0.91 |
| fast progressors | 0.77 | 0.07 | -0.11 – 0.97 |
| slow progressors | 0.76 | 0.01 | 0.29 – 0.93 |
| Knee extension × $T_{2mTHIGH}$ | 0.36 | 0.18 | -0.18 – 0.74 |
| fast progressors | 0.18 | 0.74 | -0.74 – 0.84 |
| slow progressors | 0.71 | 0.03 | 0.09 – 0.93 |
| Plantar flexion × $fRMA_{TRICEPSSUR}$ | 0.78 | <0.001 | 0.47 – 0.92 |
| fast progressors | 0.86 | 0.06 | -0.09 – 0.99 |
| slow progressors | 0.86 | <0.001 | 0.53 – 0.96 |
| Plantar flexion × $T_{2mCALF}$ | 0.57 | 0.03 | 0.08 – 0.84 |
| fast progressors | 0.85 | 0.07 | -0.12 – 0-99 |
| slow progressors | 0.30 | 0.41 | -0.41 – 0.78 |
| Grip × $VOL_{HAND}$ | 0.71 | 0.004 | 0.29 – 0.90 |
| fast progressors | 0.52 | 0.48 | -0.88 – 0.99 |

| | | | |
|---|---|---|---|
| slow progressors | 0.80 | 0.005 | 0.35 – 0.95 |
| IOPI × fRMA$_{TONGUE}$ | 0.27 | 0.48 | -0.67–1.30 |
| fast progressors | n.a. | n.a. | n.a. |
| slow progressors | 0.24 | 0.57 | -0.92–1.53 |
| Jaw opening × VOL$_{PL}$ | 0.15 | 0.59 | -0.39 – 0.61 |
| fast progressors | 0.29 | 0.64 | -0.80 – 0.93 |
| slow progressors | 0.16 | 0.65 | -0.52 – 0.72 |

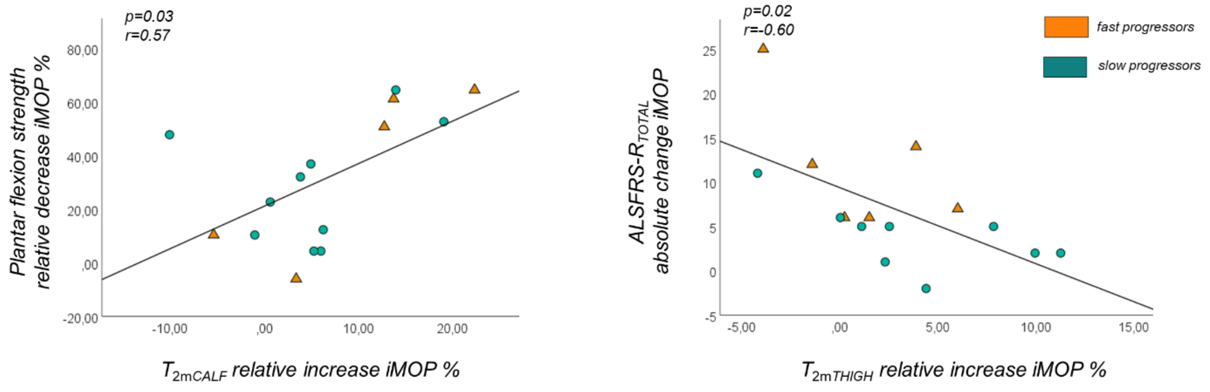


**Supplementary Figure 3**. Correlation of longitudinal water T2 changes vs. clinical parameters over individual maximum observation period (iMOP). Left panel: At calf level, the relative increase in $T_{2mCALF}$ significantly correlated with the decrease of plantar flexion strength over time (r=0.57, p=0.03), without differences between fast and slow progressors (z=.1.37, p=0.17). Right panel: At thigh level, the relative increase of $T_{2mTHIGH}$ over iMOP correlated with an increase in functional rating scale scores (ALSFRS-R$_{TOTAL}$), without differences between fast and slow progressors (z=0.22, p=0.83). Note: ALSFRS-R$_{TOTAL}$: amyotrophic lateral sclerosis functional rating scale revised total score; $T_{2mCALF}$: water T2m at calf level; $T_{2mTHIGH}$: water T2 at thigh level.

#### ***Progressive loss of fRMA and muscle volume correlate with disease severity***

##### ***Head and neck***

At head-neck level, the relative decrease of overall tongue $fRMA_{TONGUE}$ significantly correlated with the decrease of the subscale $ALSFRS_{BULBAR-SS}$ over time (rho=0.78, p=0.003, **Supplementary Table 3**). Comparison of correlations between fast progressors and slow progressors could not be performed due to loss of follow-up of 3 fast progressors.

Furthermore, the relative volume decrease of the bilateral pterygoideus lateralis muscles correlated also with the decrease of the subscale $ALSFRS_{BULBAR-SS}$ over the iMOP (rho=045, p=0.04, **Supplementary Table 3**). Correlations did not significantly differ between fast (r=0.44, p=0.46) and slow progressors (rho=0.34, p=0.35; z=0.15, p=0.88)

No significant correlations between quantitative MRI parameters and myometry assessments were observed at head-neck level.
